# PheWAS analysis on large-scale biobank data with PheTK

**DOI:** 10.1101/2024.02.12.24302720

**Authors:** Tam C. Tran, David J. Schlueter, Chenjie Zeng, Huan Mo, Robert J. Carroll, Joshua C. Denny

**Affiliations:** National Human Genome Research Institute, National Institutes of Health, Bethesda, MD; University of Toronto, Ontario, Canada; Vanderbilt University School of Medicine, Nashville, TN; *All of Us* Research Program, National Institutes of Health, Bethesda, MD

## Abstract

**Summary:** With the rapid growth of genetic data linked to electronic health record data in huge cohorts, large-scale phenome-wide association study (PheWAS), have become powerful discovery tools in biomedical research. PheWAS is an analysis method to study phenotype associations utilizing longitudinal electronic health record (EHR) data. Previous PheWAS packages were developed mostly in the days of smaller biobanks and with earlier PheWAS approaches. PheTK was designed to simplify analysis and efficiently handle biobank-scale data. PheTK uses multithreading and supports a full PheWAS workflow including extraction of data from OMOP databases and Hail matrix tables as well as PheWAS analysis for both phecode version 1.2 and phecodeX. Benchmarking results showed PheTK took 64% less time than the R PheWAS package to complete the same workflow. PheTK can be run locally or on cloud platforms such as the *All of Us* Researcher Workbench (*All of Us*) or the UK Biobank (UKB) Research Analysis Platform (RAP).

**Availability and implementation:** The PheTK package is freely available on the Python Package Index (PyPi) and on GitHub under GNU Public License (GPL-3) at https://github.com/nhgritctran/PheTK. It is implemented in Python and platform independent. The demonstration workspace for *All of Us* will be made available in the future as a featured workspace.

## INTRODUCTION

Since the introduction of PheWAS over a decade ago, it has become a powerful technique to study the association of genetic variants and phenotypic variations across human populations (Bastarache, et al., 2022; Denny, et al., 2010). A PubMed search in December 2023, using the search terms of “PheWAS” or “phenome-wide association” in Title/Abstract field, returned 679 publications, from only 3 publications in 2010 to 159 publications in 2023, conducted in many biobanks and countries, including *All of Us* (United States), UK Biobank, BioBank Japan, and many others (Dofuku, et al., 2023; Millwood, et al., 2016; Rao, et al., 2018; Schlueter, et al., 2023). PheWAS has been used to address different scientific objectives, such as refining results in conjunction with genome-wide association studies (GWAS), studying comorbidities and precision subsets of disease, and evaluating potential repurposing of medications and predicting adverse drug reactions (Bastarache, et al., 2022).

The rapid development in sequencing technology, computational power, and cloud infrastructure has led to significant increase in genetic and phenotype data in major biobanks worldwide. In 2023, UKB released whole genome sequencing (WGS) data on 500,000 participants linked to decades of health record data (Callaway, 2023), and *All of Us* increased their WGS data to ~245,000 participants and 40+ years of health record data (Ginsburg, et al., 2023). This enormous amount of available data can create a barrier for researchers due to the potential costs and time-consuming nature of analysis. Moreover, data are often organized in different formats, e.g., VCF, Plink or Hail formats for genetic data and Observational Medical Outcome Partnership (OMOP) for health data, which require specific technical knowledge to utilize effectively.

Many packages have been developed for PheWAS, such as the R PheWAS package (Carroll, et al., 2014), PHESANT (Millard, et al., 2018), or pyPheWAS (Kerley, et al., 2022). However, these packages face certain limitations, including utilizing single-threaded computational approaches, lacking support for common data models, tools to assist with major steps in a PheWAS workflow, and/or lacking newer phecodeX implementation, which expands the number of phecodes to >3600. To address these limitations, we developed PheTK, an efficient Python package supporting end-to-end standard PheWAS analyses.

## METHODS

### General PheWAS workflow and PheTK main functionalities

PheTK was developed to handle large datasets quickly, utilizing specific optimizations and multithreading. Moreover, PheTK supports routine procedures such as extraction of variant data from Hail Matrix tables, a scalable data storage format (Hail, 2008), and phenotype data from OMOP databases. The package is currently structured into four main modules – Cohort, Phecode, PheWAS and Plot – which assist users with various steps of a complete PheWAS analysis pipeline (**Figure 1; Table S1**). Users have the option to use their own data or utilize PheTK’s modules for these tasks. In addition, we created Demo module to help new users familiar with PheWAS.

**Figure 1.**
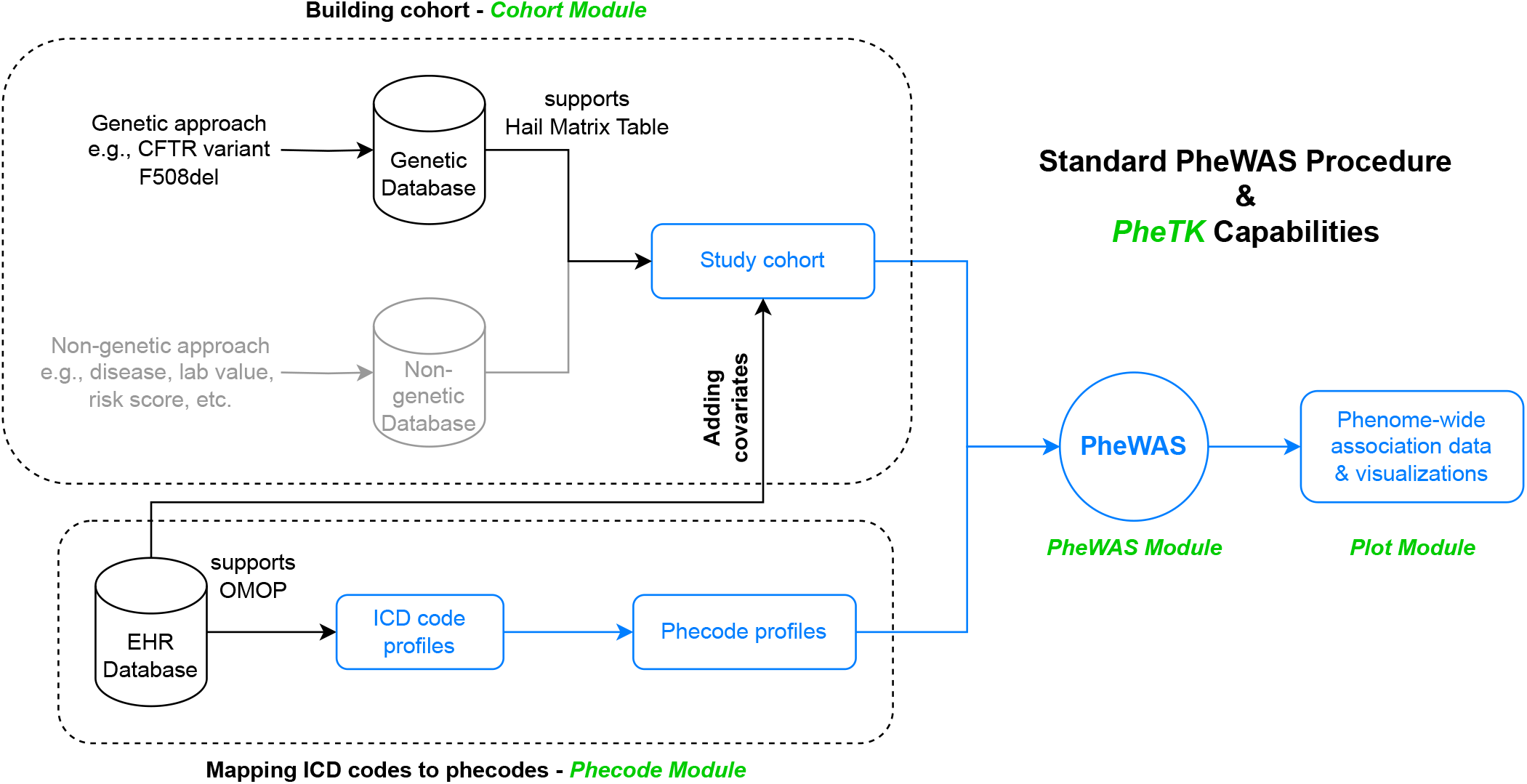
Standard PheWAS procedure and PheTK capabilities. Components in black and blue colors are supported by PheTK while components in gray are not supported. The blue components were assessed in benchmark test against the R PheWAS package. PheTK module names are in green.

### Building a cohort

A typical PheWAS study starts with a genetic variant. The Cohort module generates a cohort from genetic data stored in Hail matrix tables, a widely used data storage format for biobank level databases including *All of Us*. This function accepts user input containing variant genomic position, allele change and path to the Hail matrix table, then returns a case/control cohort with participants possessing the specific genotypes. In addition, the Cohort module supports generation of common covariates, such as age at last event, sex at birth, EHR length, and others from OMOP databases, which is widely adopted in the field of health informatics and usually requires knowledge of SQL to query data. PheTK uses a combination of SQL queries and subsequent data processing to obtain covariate data based on user inputs.

### Phecode mapping

A core component of a PheWAS analysis workflow is mapping International Classification of Disease (ICD) codes to phecodes. The Phecode module retrieves ICD codes (supporting ICD versions 9-CM, 10, and 10-CM) from compatible OMOP databases and subsequently maps and aggregates ICD codes to phecodes, currently supporting both version 1.2 and the recently released phecodeX 1.0 (Shuey, et al., 2023). Users can also provide their own ICD code data, or their own ICD to phecode map, as an input into the phecode mapping function within the Phecode module to generate phecode counts.

### Statistical analysis

With PheTK, users can customize settings for their analysis, such as the minimum phecode count required to define a case, the minimum number of cases to study, and whether to apply phecode exclusion criteria, as in the R PheWAS package (Carroll, et al., 2014). The PheWAS module accepts cohort data and phenotype data either generated by the Cohort and Phecode modules or provided directly by users as inputs. For each phecode, the input data is processed and divided into case/control groups which are then used in logistic regression. Parallel processing through multithreading allows for efficient execution of these regressions. The module provides summary statistics for each phecode, including the number of cases and controls, p-value, beta coefficient, confidence intervals, odds ratio, and Bonferroni correction threshold of the independent variable of interest.

### Visualization

The Plot module offers support for Manhattan and volcano plots for PheWAS result visualization. This module comes with multiple customization options, such as labeling specific phenotypes or certain categories, labeling by order of p-values or beta coefficients, changing label colors, marker sizes, figure resolution, and more. This module was built on top of Python matplotlib library (Hunter, 2007), which could be used for further customizations.

### Application examples and performance benchmarking

To demonstrate the package, we performed a PheWAS for one genetic and one non-genetic phenotype in *All of Us*. Our evaluation included a benchmarking test comparing PheTK with the R PheWAS package, which also supports multithreading. We assessed PheTK’s performance across various working environments, including *All of Us*, UKB RAP, and a laptop, utilizing both phecode 1.2 and phecodeX to demonstrate its flexibility.

A genetic cohort consisting of 181,776 participants was selected from the *All of Us* database based on the presence or absence of the most common pathogenic variant causing cystic fibrosis, *CFTR* F508del or rs113993960 (Lukacs and Verkman, 2012). This variant was identified as chr7-117559590-ATCT-A using *All of Us* genomic data browser. Phenotype profiles of all cohort participants were generated, and PheWAS was performed using an additive model adjusted for age at last event, sex at birth, and 10 genetic principal components (PC) provided by *All of Us*.

In addition, a uric acid study cohort of 9,170 participants was created to demonstrate a PheWAS analysis with a non-genetic cohort using a continuous variable. Urate (mass/volume) in serum or plasma measurements with proper unit and values were retrieved from the *All of Us* OMOP database using custom SQL queries. Participants with at least three urate measurements were selected and all-time average urate values were calculated and used as variable of interest. PheWAS was performed adjusted for age at last event, sex at birth, and 10 PCs. To demonstrate PheTK’s compatibility with different phecode versions, phecode 1.2 was used for the *CFTR* PheWAS and phecodeX was used for the uric acid PheWAS.

We used the *CFTR* study as a test case for benchmarking PheTK performance compared to the R PheWAS package. The test environment, setup on the *All of Us*, was a standard virtual machine (VM) with 96 CPUs and 360 GB RAM. Both packages were set to use 64 CPUs to ensure sufficient computing power for background tasks, if any. We benchmarked the same processes, which included mapping ICD codes to phecodes, subsequent case/control processing for each phecode, and logistic regressions of qualified phecodes (minimum case number = 20, minimum phecode count = 2; **Figure 1**). All component times were combined to get the total runtime of each package.

To demonstrate package compatibility with other platforms, we performed a similar *CFTR* F508del PheWAS study on UKB Research Analysis Platform (RAP). We generated Hail matrix table from UKB whole exome sequencing (WES) pVCF data and used the Cohort module to create F508del cohort from the matrix table. We used phecode 1.2 for this cohort of 469,007 participants adjusted for age at recruitment, sex, and 10 PCs. VM configuration was 96 CPUs and 375 GB RAM with number of CPUs used set to 64 in PheTK settings. Finally, we ran PheWAS analysis of the same UKB *CFTR* cohort on a MacBook Pro M2 Pro 12 CPU cores 32GB RAM using phecodeX, which had nearly twice as many phecodes than phecode 1.2.

## RESULTS AND DISCUSSION

### Application examples

PheWAS is a powerful yet complex discovery tool with multiple steps that deal with different types of data in different formats. PheTK can accelerate scientific discoveries by providing tools for quickly tackling common technical tasks with support for common data formats. As expected, the *CFTR* PheWAS demonstration PheWAS in *All of Us* found highly significant associations with cystic fibrosis (p-value = 1.3 × 10^−51^, OR = 26.8) and other related phenotypes (**Figure S1A-B**). Every step was performed exclusively by calling PheTK modules without the need to write custom scripts, with no knowledge of Hail, OMOP, or SQL required. Total runtime was less than 15 minutes, from generating a cohort to result visualization.

The UKB *CFTR* PheWAS on RAP completed successfully in 11.9 minutes using phecode 1.2. While having a significantly larger cohort size than the *All of Us* test cohort (469,007 vs 181,776 participants), there were only 1,423/1,624 phecodes that met count thresholds versus 1,765/1,816 phecodes in the *All of Us CFTR* PheWAS. As with the *All of Us* analysis, cystic fibrosis (p-value = 1.9 × 10^−10^, OR = 10.38) was associated with *CFTR* F508del (**Figure S1C**).

In the non-genetic approach demonstration, the uric acid PheWAS with phecodeX workflow executed by PheTK modules took 5 minutes to complete with expected associations found between high plasma/serum urate concentration and gout (p-value = 2.2 × 10^−184^, OR = 1.83) and chronic kidney disease (p-value = 2.2 × 10^−165^, OR = 1.64). These results are visualized using the Manhattan and volcano plotting options (**Figure 2**). Except for custom SQL queries to generate the cohort from lab measurement data, all other steps were performed by calling PheTK modules.

**Figure 2.**
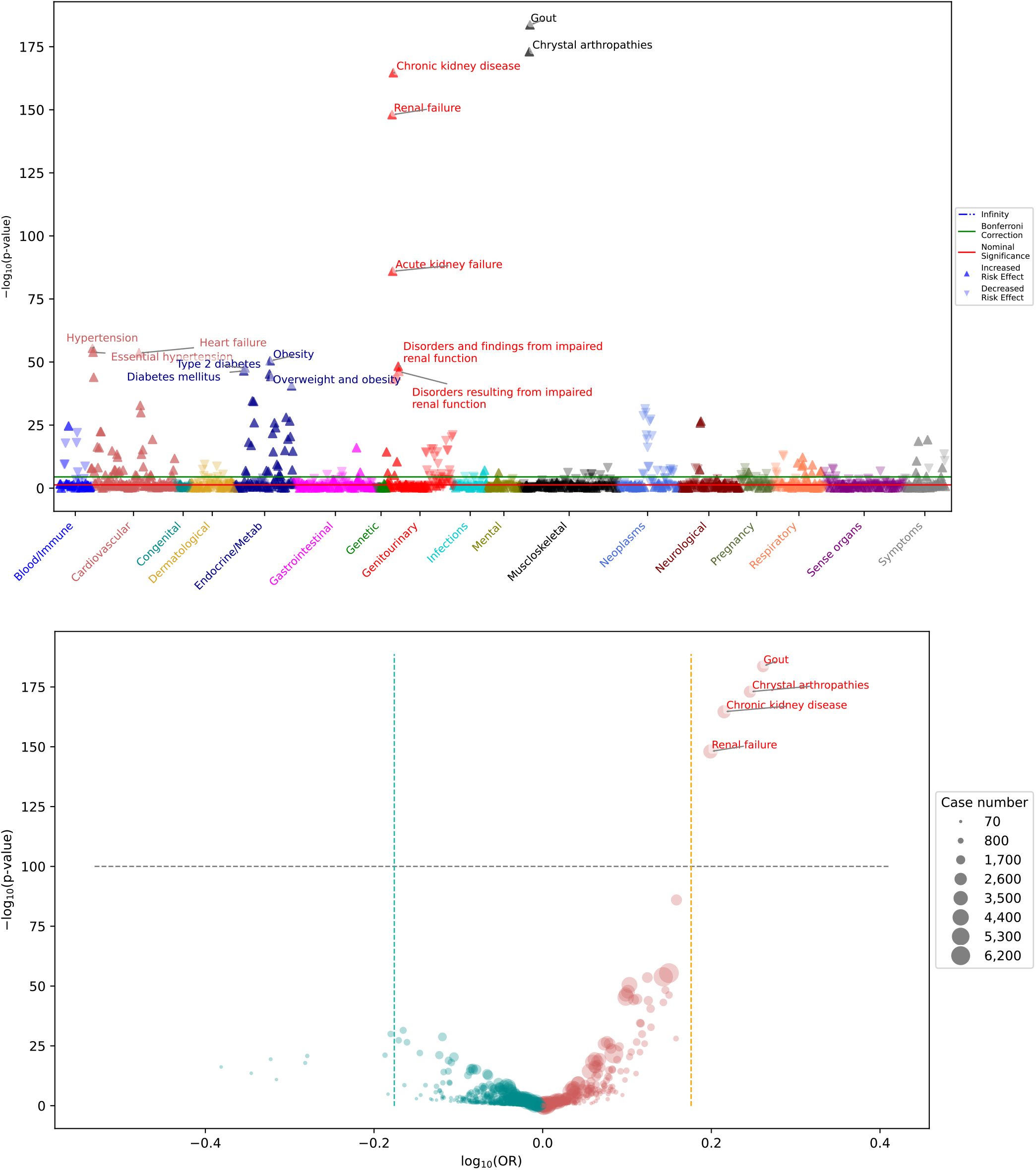
Uric acid PheWAS analysis with phecodeX. **(A)** Manhattan plot displays significant positive associations between uric acid level and various phenotypes, including gout, chronic kidney disease, and others. **(B)** Volcano plot highlights the significant phenotypes with large effect sizes, such as gout, crystal arthropathies, chronic kidney disease, and renal failure.

### Benchmarking and other performance tests

In the benchmark test, PheTK took 64% less time than the R PheWAS package to finish the same workflow (9.9 minutes compared to 27.3 minutes). Based on the cost of the test environment of $4.65 per hour, the test workflow cost $0.77 and $2.12 for PheTK and the R PheWAS, respectively. It should be noted that this VM configuration was chosen to avoid bottlenecks due to insufficient computing resources, and cheaper configurations could also be used.

We then tested PheTK performance in limited computing resources of a MacBook Pro M2 Pro laptop by running PheWAS regressions with phecodeX using the largest dataset in this publication (469,007 UKB *CFTR* cohort). PheTK successfully completed the analysis in ~81 minutes with 2192/2721 phecodes tested, with cystic fibrosis as most significant phenotype (**Figure S1D**).

## CONCLUSION

PheTK provides scalable performance across large datasets on different platforms. The software can be installed via PyPi and run on any Python environment, including cloud VMs, institutional HPCs, or local workstations or laptops. While applicable to any ICD and genetic data, PheTK has been optimized to simplify the steps for *All of Us* and other biobanks with similar data structures. We plan to add support for other platforms, standard data formats, and additional analysis methods over time.

## Data Availability

All data produced in the present work are contained in the manuscript

## ACKNOWLEDGEMENTS

We would like to express our gratitude to internal and external collaborators, including all Denny lab members, the Cohort Analytics Core at the National Human Genome Research Institute (NHGRI), and Dr. Jason Karnes and Dr. Kiana Martinez from University of Arizona, who kindly tested the package and provided essential feedback, greatly improving this work. We would like to extend our gratitude to Dr. Tracey Ferrara (NHGRI) for helping with manuscript review and administrative assistance.

---

**Figure S1.**
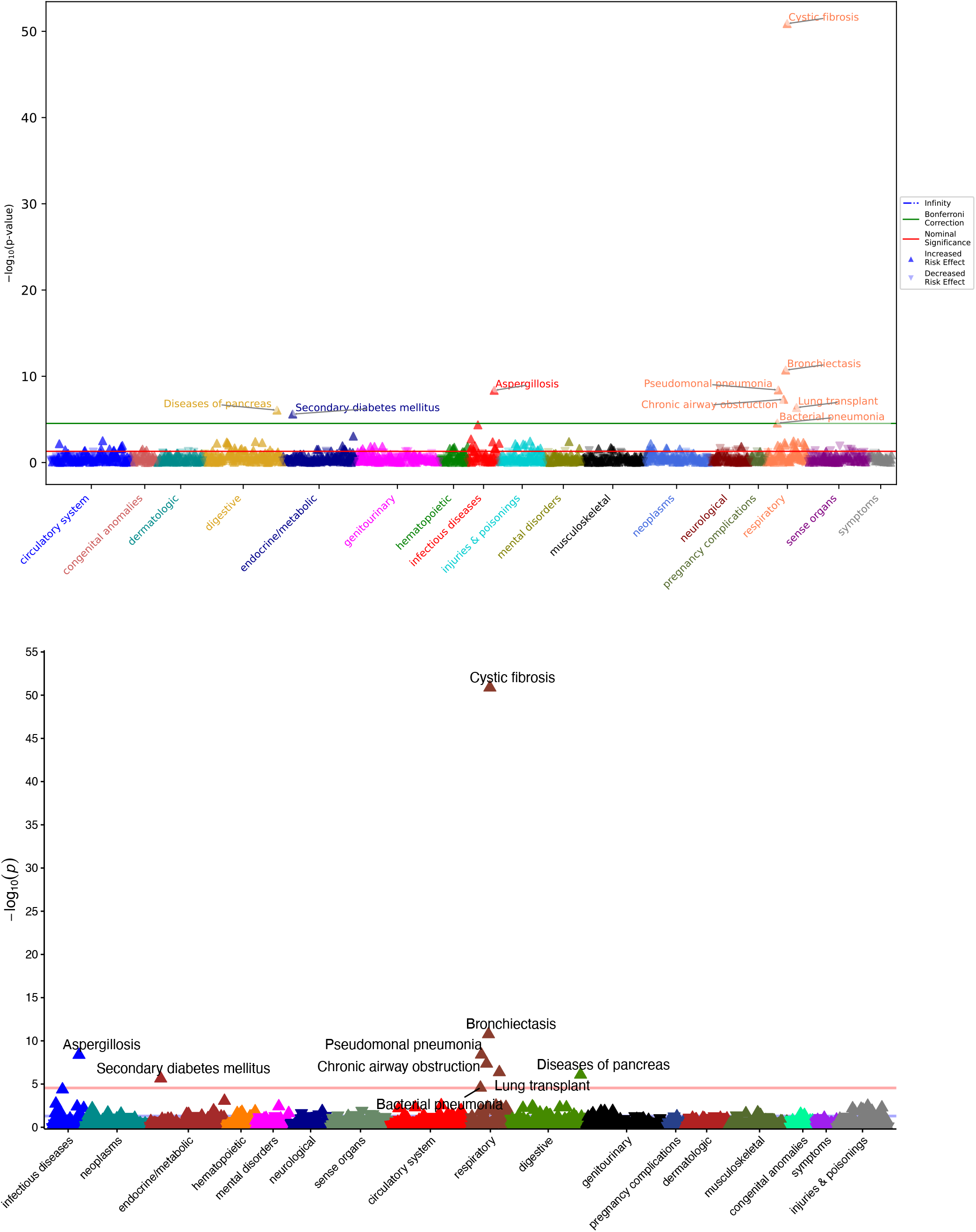
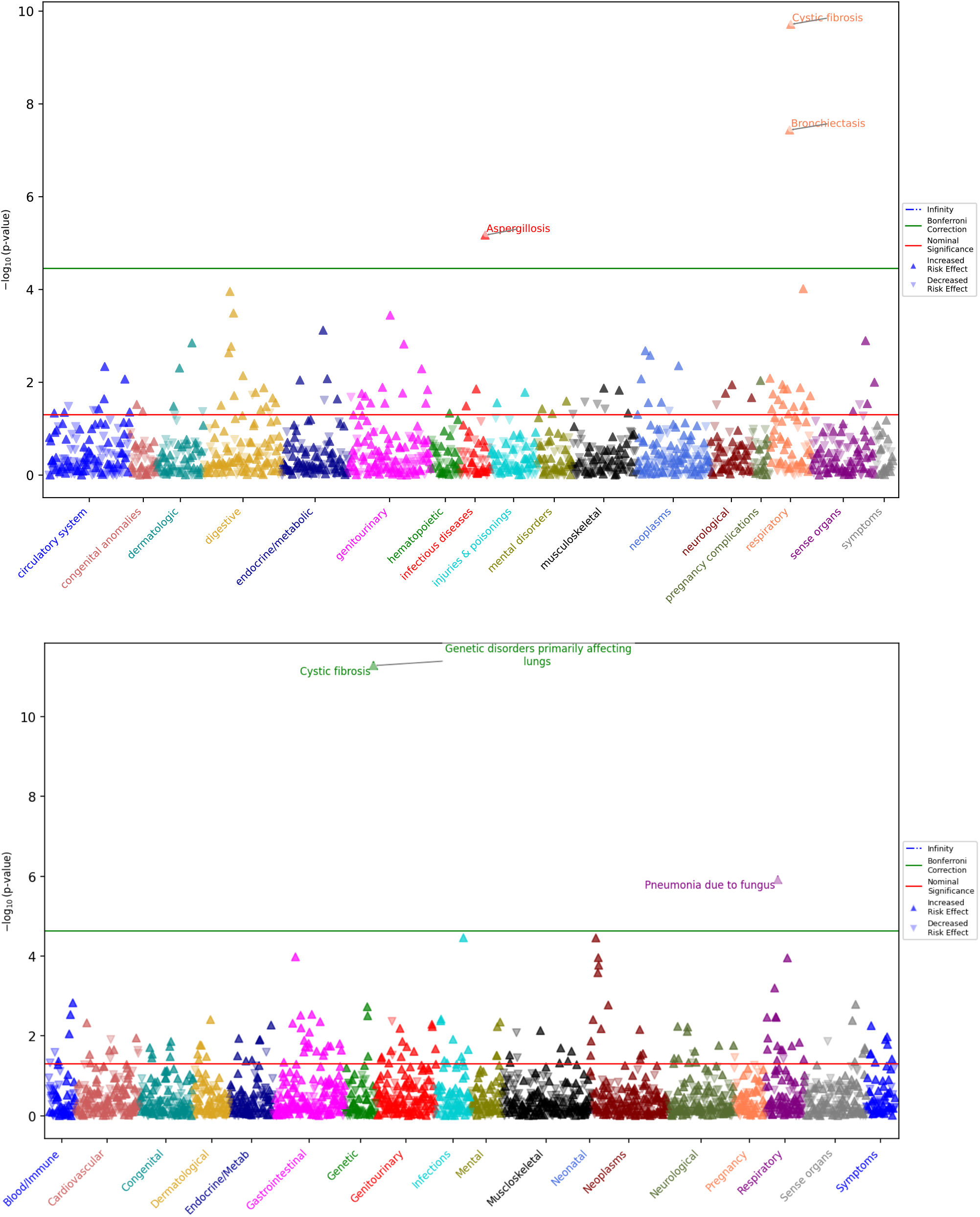
CFTR variant F508del PheWAS analysis. PheWAS results using phecode 1.2 on All of Us with PheTK **(A)** and the R PheWAS package **(B)**, PheWAS with phecode 1.2 on UKB RAP with PheTK **(C)**, and PheWAS with phecodeX on a MacBook Pro with PheTK **(D)**. All results show significant association of cystic fibrosis and F508del variant.

**Table S1.** PheTK module descriptions.

| PheTK modules | Usage in PheWAS workflow | Notes |
| --- | --- | --- |
| **Cohort** | - Generates cohorts from phenotypes of variant of interest, e.g., F508del (chr7-117559590-ATCT-A) cohort with 0/1 genotype as cases and 0/0 controls.<br>- Extracts data for covariates commonly used in PheWAS, including age at last event, sex at birth, EHR length, genetic principal components, etc., from EHR data. | - Cohort generation from genetic data stored in Hail matrix table format is supported. This feature works with Hail matrix tables on any platform.<br>- Data extraction from OMOP standard tables in Google BigQuery is supported. Database must have “person”, “condition_occurrence”, “observation”, “death”, and “concept” tables. This feature is optimized for *All of Us* and will require further customizations for other platforms. |
| **Phecode** | - Counts ICD codes from EHR data and maps ICD codes to phecode 1.2 or phecodeX, using official PheWAS Catalog or user provided mapping tables. | - Extracting ICD codes from OMOP data in BigQuery having required tables mentioned above is supported.<br>- Mapping ICD codes to phecodes works on any platform with user provided ICD code data. |
| **PheWAS** | - Runs PheWAS analysis. | - Currently logistic regression is supported, utilizing Logit function from python statsmodels library. |
| **Plot** | - Visualizes PheWAS results. | - Manhattan plot and volcano plot are supported. Both plotting functions were developed using python matplotlib library. |
| **Demo** | - Includes a quick 1-minute demo to help new users familiar with PheWAS analysis. |  |

